# Multimodal longitudinal profiling of bacterial composition, viable burden, and spatial distribution in recessive dystrophic epidermolysis bullosa wounds

**DOI:** 10.64898/2026.07.22.26358692

**Authors:** Katja Holmgren, Enikö Sonkoly, Sigrid Lundgren, Torborg Hoppe, Bo Nilson, Emma Hundevadt, Holger Brüggemann, Artur Schmidtchen, Karl Wallblom

## Abstract

Recessive dystrophic epidermolysis bullosa (RDEB) is an inherited disorder characterized by recurrent wounds and frequent bacterial colonization. Clinically applicable methods that provide information on bacterial composition, viable burden, and spatial distribution are needed to support wound assessment and therapeutic evaluation. In this exploratory four-week longitudinal study, standard-of-care-treated wounds from five patients with RDEB were examined using autofluorescence imaging, quantitative and chromogenic culture, MALDI-TOF MS, 16S rRNA and staphylococcal *tuf2* amplicon sequencing, and spatial Bactogram analysis. Sequencing showed that staphylococci dominated 22/23 communities, with *Staphylococcus aureus* being the most abundant staphylococcal species (21/23). Culture likewise identified *S. aureus* as the most frequent species in all 14 swab samples. Aerobic bacterial burden showed no association with open wound area and remained high in several contracting wounds. The Bactogram provided a spatial representation of viable bacterial growth and, when combined with chromogenic agar, reflected the predominance of *S. aureus* identified by sequencing, MALDI-TOF MS, and quantitative culture. Together, these complementary methods captured the dynamic nature of RDEB wounds through heterogeneous healing trajectories and longitudinal variation in bacterial burden, while consistently identifying a low-diversity, *S. aureus*-dominated microbiota. Bactogram spatial imprinting and autofluorescence imaging warrant further evaluation as minimally invasive tools for longitudinal EB assessment.

## Introduction

Epidermolysis bullosa (EB) is a group of inherited disorders characterized by fragility and blistering of the skin and other stratified epithelia (Has et al., 2020). Dystrophic EB (DEB) is caused by pathogenic variants in *COL7A1*, encoding type VII collagen, a key component of anchoring fibrils at the dermal–epidermal junction (Cianfarani et al., 2017; Has et al., 2020). The recessive form, RDEB, includes severe phenotypes with markedly reduced or absent functional collagen VII, leading to widespread, recurrent wounding and considerable morbidity, including pain, scarring, and an increased risk of infection, sepsis, and cutaneous squamous cell carcinoma (Cianfarani et al., 2017; Fine et al., 2008; Foll et al., 2018; Mellerio, 2010; Nystrom et al., 2021). Even with recent advances in local corrective gene therapies, there is still no cure, with patients requiring lifelong wound monitoring and care (Bardhan et al., 2020; Guide et al., 2022; Tang et al., 2025).

The microbiology of EB wounds has been investigated using both culture-based and sequencing-based approaches. Culture-based studies have repeatedly identified *Staphylococcus aureus* as a prominent wound organism in EB, often alongside other wound-associated bacteria such as *Pseudomonas aeruginosa*, streptococci, and corynebacteria (Brandling-Bennett and Morel, 2010; Fuentes et al., 2023; Levin et al., 2021; Singer et al., 2018). Culture-independent sequencing studies, including RDEB-specific and broader EB/DEB cohorts, have further shown that EB-involved skin and wounds often exhibit reduced microbial diversity and enrichment of *Staphylococcus* species, consistent with disease-associated dysbiosis (Bar et al., 2021; Fuentes et al., 2018; Horev et al., 2023; Reimer-Taschenbrecker et al., 2022). These findings are clinically relevant because impaired barrier function, repeated cycles of wounding and repair, and an altered antibacterial host defense place patients with DEB at high risk of bacterial colonization and infection, which contribute to morbidity, delayed wound healing, and reduced quality of life (Foll et al., 2018; Fuentes et al., 2023; Huitema et al., 2021; Nystrom et al., 2018). Accurate assessment of composition, viable burden, and spatial distribution in DEB wounds is therefore important for clinical wound evaluation and for assessing therapeutic interventions.

In clinical practice, bacterial assessment in EB still largely relies on visual inspection and clinical symptoms, with conventional swab culture used when necessary to identify potential pathogens (Denyer et al., 2017). However, these approaches have important limitations. Generally, wounds may harbor high bacterial loads without overt clinical signs of infection, while biofilm formation and spatial heterogeneity can complicate sampling and interpretation (Huitema et al., 2021; Kirkup, 2015; Le et al., 2021; van der Kooi-Pol et al., 2012). Conventional culture detects only organisms that are viable and grow under the chosen conditions, although it does allow the viable load to be quantified (Misic et al., 2014). Standard amplicon sequencing approaches, by contrast, provide broader community information, but do not reliably distinguish viable from nonviable bacteria and, unless paired with an independent measure of total bacterial load, primarily provide compositional data (Misic et al., 2014; Ren et al., 2021; Tettamanti Boshier et al., 2020). Moreover, a conventional swab samples only a limited region and may not capture spatially heterogeneous bacterial colonization across the wound and surrounding skin. This is particularly relevant in RDEB, where repeated sampling may cause discomfort and additional trauma to fragile skin.

In the present exploratory study, we applied a comprehensive multimodal microbiological approach to wounds from five patients with RDEB, sampled repeatedly over four weeks. We compared culture-based methods, including quantitative colony-forming unit (CFU) analysis and matrix-assisted laser desorption/ionization time-of-flight mass spectrometry (MALDI-TOF MS) species identification, with molecular analyses based on 16S rRNA and staphylococcal *tuf2* gene profiling, the latter providing improved species-level resolution of staphylococci (Feidenhansl et al., 2024). We additionally compared two clinically applicable approaches for spatial bacterial assessment: noninvasive autofluorescence imaging (Le et al., 2021) and Bactogram spatial imprint culturing (Wallblom et al., 2024a), which retains the spatial distribution of viable bacteria across the wound and the surrounding skin. To our knowledge, neither spatial approach has previously been evaluated in RDEB wounds. Our aims were to determine the complementary information provided by these modalities and their concordance, and to assess how viable bacterial burden relates to clinical wound status over time.

## Methods

### Study design

This is an exploratory microbiological analysis of standard-of-care control wounds sampled during a non-randomized, open-label, single-arm phase I study of the immunomodulatory peptide TCP-25 in DEB. The trial design and safety results have been reported in full in Wallblom et al. (2025). The present analysis is restricted to standard-of-care reference wounds and their corresponding healed skin, thereby allowing the natural wound microbiology of DEB to be characterized without confounding effects from the investigational treatment.

Briefly, five patients with RDEB were enrolled at the Dermatology Clinic, Skåne University Hospital, Lund, and at Clinical Trial Consultants AB, Uppsala, Sweden (recruited from Uppsala University Hospital, Uppsala, Sweden), between November 2023 and March 2024. Each patient had two wound areas treated with TCP-25 alongside a control wound area receiving standard wound care only. Standard wound care was defined as the regular wound treatment of DEB at the clinics and consistent with expert recommendations (Denyer et al., 2017), but the choice of dressing was limited to a silicone-based polyurethane wound dressing (Mepilex Lite, Mölnlycke, Gothenburg, Sweden). In practice, all study wound areas were gently cleaned with physiological saline solution at all dressing changes, according to established routines for EB wounds. Patients were followed from Day 1 to Day 29, with an end-of-study phone call on Day 32. Figure 1 summarizes the timing of clinical imaging, autofluorescence imaging, swab collection, dressing collection, and Bactogram sampling. Further details regarding inclusion and exclusion criteria, study restrictions, and study design are available in Wallblom et al. (2025).

**Figure 1.**
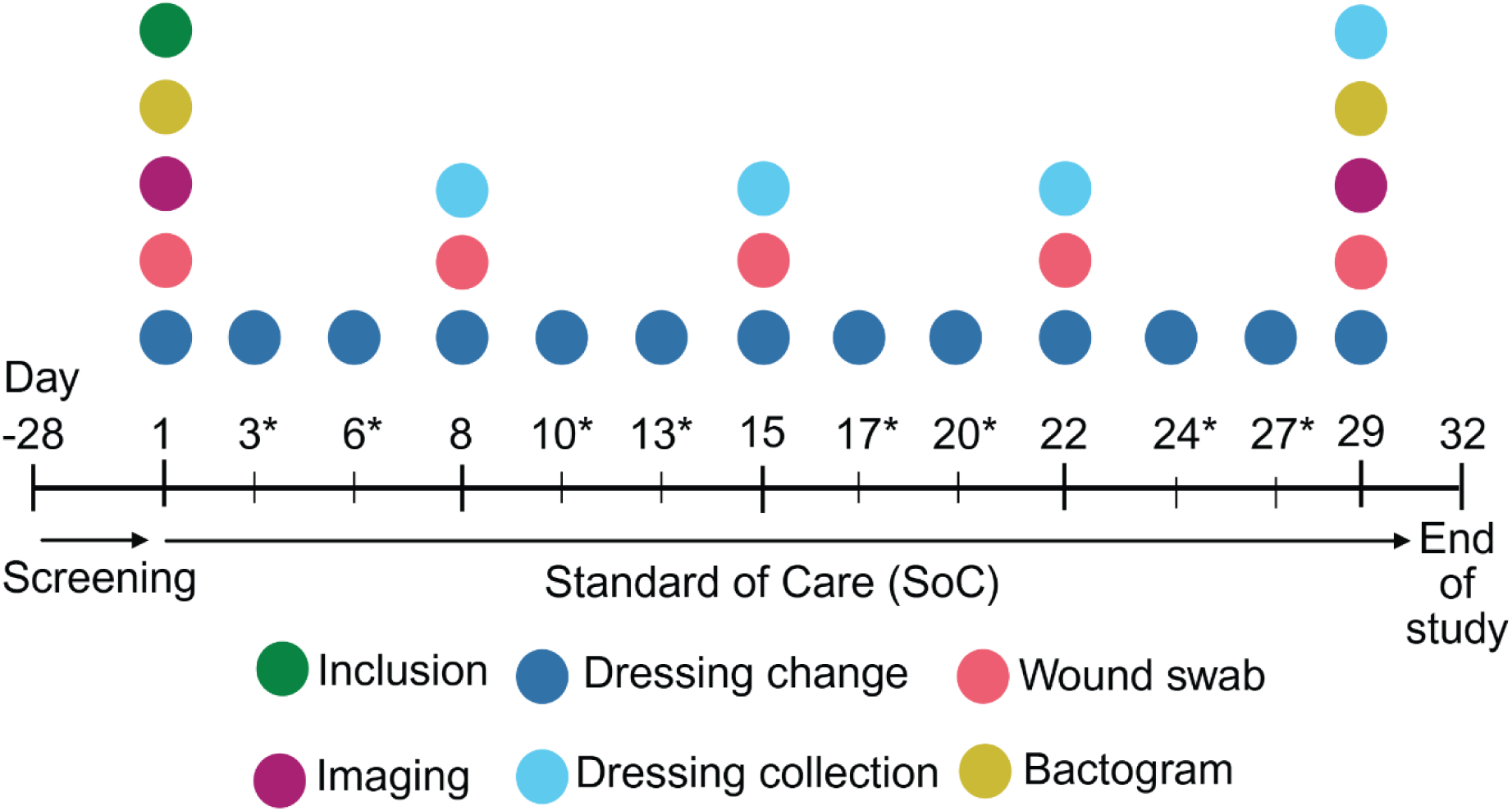
Schedule of assessments. After screening (Day −28 to Day 1), patients were followed according to standard of care from Day 1 to Day 29, with an end-of-study phone call on Day 32. Colored markers indicate study events (inclusion, dressing change, wound swab, imaging, dressing collection, Bactogram). *Asterisks denote at-home dressing changes done by the patient or caregiver, preferably on these days.

### Clinical photography and autofluorescence imaging

As previously reported (Wallblom et al., 2025), the open wound area was measured by laser-assisted planimetry using a SilhouetteStar (ARANZmedical, Christchurch, New Zealand) camera system and the Silhouette Connect program (ARANZmedical, Christchurch, New Zealand) at the time of the screening and on Days 1, 8, 15, 22, and 29. Wounds were photographed after cleaning.

High-quality clinical photographs were obtained after cleaning at the first (Day 1) and final (Day 29) study visits using a DSLR camera (EOS Rebel T7i, Canon, Tokyo, Japan) fitted with a 60 mm f/2.8 macro lens (EF-S 60 mm, Canon, Tokyo, Japan). Standardized lighting was provided by a ring flash system (Twin Flash, Canfield, Fairfield, NJ, USA), and fixed camera settings were used throughout. A disposable ruler with color references was positioned slightly above the wound as a scale and color standard. To ensure reproducible image acquisition, the distance to the wound was estimated by aligning the two range-finder laser dots to be approximately 7 cm apart, as measured on the ruler. This translated into an approximate distance of 65 cm between the camera and the wound. This standardized photography protocol was based on the method previously described by Wallblom et al. (2024b).

Autofluorescence imaging of bacteria was performed using a handheld device (MolecuLight i:X, MolecuLight Inc., Toronto, Canada) before wound cleaning and Bactogram collection. Images were acquired according to the manufacturer’s instructions, using dark drapes and violet excitation light at 405 nm to detect endogenous bacterial autofluorescence associated with an elevated bacterial burden (>10⁴ CFU/g) (Le et al., 2021). Fluorescence signals were interpreted qualitatively according to the manufacturer’s instructions and used as a complementary, noninvasive spatial indicator of bacterial presence.

### Swab collection and handling

Wounds were swabbed at the time points indicated in Figure 1. At each time point, two swabs were collected from each patient: a cotton swab (Selefa, Stockholm, Sweden) and a FLOQswab (Copan, Brescia, Italy). Both swabs were taken from approximately the same central area of the wound by applying steady pressure and rotating the swab back and forth five times. When the wound had re-epithelialized, the corresponding area of newly healed skin was sampled. Swabs were collected after standard wound cleaning with physiological saline to remove contaminants and superficial, less clinically relevant bacteria (Kelly, 2003).

The FLOQswab was stored in eNAT (Copan, Brescia, Italy) for microbiome analysis because eNAT lyses bacteria and preserves microbial nucleic acids (Young et al., 2020). The cotton swab was placed directly into a 1.5 mL tube containing 0.5 mL of ice-cold phosphate-buffered saline (PBS) and kept on ice until analysis. The tubes were vortexed to dislodge bacteria from the swab before analysis. A previous study by Lundgren et al. showed that storage under these conditions for 24 hours does not significantly affect the viability of common skin bacteria (Lundgren et al., 2025).

### Dressing fluid extraction

All study wounds were covered with a standardized 5 × 10 cm silicone-based polyurethane dressing (Mepilex Lite, Mölnlycke, Gothenburg, Sweden), which was changed according to the schedule shown in Figure 1. At the beginning of the specified study visits, used dressings were collected in a sterile 15-cm Petri dish and placed directly on ice. Each dressing was first cut in half, and each half was transferred to a 50 mL tube containing 25 mL of ice-cold 10 mM Tris (pH 7.4; 1 mL/cm^2^). The tube was vortexed for 5 minutes. The dressing halves were then transferred to a 20 mL syringe, compressed to recover any remaining fluid back into the tube, and discarded.

### Identification of aerobic cultivable species using MALDI-TOF MS

Swab and dressing fluid samples from specified time points (Days 1, 15, and 29 for swabs; Days 15 and 29 for dressings) were streaked onto blood agar and incubated in aerobic conditions (5% CO₂ at 37 °C) overnight (16 hours). Blood agar was chosen because it is routinely used in clinical microbiology laboratories and supports the growth of the most common clinically relevant pathogens (Jorgensen et al., 2015). To obtain a representative overview of the major cultivable bacterial species, six colonies were selected from each plate. When distinct colony morphologies were present, at least one colony of each type was chosen in proportion to its relative abundance. When colony morphology was uniform, colonies were randomly selected.

Colonies were prepared on stainless steel MALDI target plates using the manufacturer’s extended direct transfer protocol (Bruker Daltonik GmbH, Bremen, Germany). Mass spectra were acquired with a Microflex LT/SH SMART MALDI-TOF MS instrument using flexControl version 3.4 (Bruker Daltonik GmbH, Bremen, Germany) in linear mode over a mass range of 2 to 20 kDa. For each sample spot, 240 summed laser shots were collected. Spectra were analyzed with MALDI Biotyper (MBT) Compass v. 4.1 and the MBT Compass Library Revision N (DB-11987, 2022) (Bruker Daltonik GmbH, Bremen, Germany). A MALDI Biotyper score of at least 2.0 was required for species-level identification according to the manufacturer’s recommendations.

### Quantification of viable aerobic cultivable bacterial load

The aerobic cultivable bacterial load was quantified from the cotton swab and dressing-fluid samples by drop-plate counting. Each sample was serially diluted in sterile PBS in seven 10- fold (1:10) steps. For each plate, six 10-µL drops of the undiluted sample and three of each dilution were deposited onto the agar surface of Todd-Hewitt plates and CHROMagar *S. aureus* plates (CHROMagar Microbiology, Paris, France), respectively. Todd-Hewitt agar is a rich medium that supports the growth of a wide range of organisms and, being more translucent than blood agar, facilitates accurate colony counting. CHROMagar *S. aureus* distinguishes presumptive *S. aureus* (pink colonies) from other bacteria (white or blue colonies). Plates were incubated at 37 °C in 5% CO₂ overnight (16 hours), and colonies were counted the following morning.

The concentration of colony-forming units (CFU/mL) was calculated from the lowest countable dilution, correcting for the dilution factor. For swabs, the total load per swab was obtained by multiplying the CFU/mL by the elution volume (0.5 mL), giving CFU/swab. For dressings, because each dressing was extracted in 1 mL of buffer per cm² of dressing, the CFU/mL of the extract corresponds directly to the bacterial load per unit area and was reported as CFU/cm² without further conversion. On CHROMagar *S. aureus*, presumptive *S. aureus* (pink) and other colonies (white or blue) were counted separately, and the *S. aureus* load was expressed as the proportion of pink colonies relative to the total colony count (pink plus non-pink).

### Culture-independent bacterial profiling

### DNA extraction

FLOQswab samples stored in eNAT were used for microbiome analysis. Prior to extraction, samples were centrifuged (8000 × g, 30 min, 4 °C) and the supernatant discarded. Pellets were lysed for 1 h with lysostaphin (0.05 mg/mL; Sigma-Aldrich, Burlington, MA, USA) and lysozyme (9.5 mg/mL; Sigma-Aldrich), and DNA was extracted with the DNeasy PowerSoil Kit (Qiagen, Hilden, Germany) following the manufacturer’s instructions. DNA concentration was measured with the Qubit dsDNA HS assay on a Qubit fluorometer (Thermo Fisher Scientific, Waltham, MA, USA).

### Amplicon PCR

The bacterial community was profiled by amplifying the 16S rRNA gene with the V1–V3 primer set (forward 5′-AGAGTTTGATCCTGGCTCAG-3′; reverse 5′- ATYACCGCGGCTGCTGGCA-3′). For staphylococcal species-level resolution, the *tuf2* gene was amplified as described previously (Feidenhansl et al., 2024) using primers *tuf2*_fw 5′- ACAGGCCGTGTTGAACGTG-3′ and *tuf2*_rev 5′-ACAGTACGTCCACCTTCACG-3′. PCR reaction mixtures were made in a total volume of 25 µL and comprised 5 µL of DNA sample, 2.5 µL AccuPrime PCR Buffer II (Invitrogen, Waltham, MA, USA), 1.5 µL of each primer (10 µM) (DNA Technology, Risskov, Denmark), 0.15 µL AccuPrime Taq DNA Polymerase High Fidelity (Invitrogen, Waltham, MA, USA), and 14.35 µL of PCR grade water. Cycling conditions were: initial denaturation at 94 °C for 2 min; 35 cycles of 94 °C for 20 s, 55 °C for 30 s, and 68 °C for 1 min; and a final elongation at 72 °C for 5 min. Products were verified on an agarose gel, purified with the Qiagen GeneRead Size Selection Kit (Qiagen, Hilden, Germany), and quantified on a NanoDrop 2000 spectrophotometer (Thermo Fisher Scientific, Waltham, MA, USA).

### Indexing and sequencing

Sample-specific indices and Illumina adapters were attached using the Nextera XT Index Kit (Illumina, San Diego, CA, USA). Index PCR contained 5 µL template, 2.5 µL of each index primer, 12.5 µL 2× KAPA HiFi HotStart ReadyMix (Roche, Basel, Switzerland), and 2.5 µL PCR-grade water, with cycling at 95 °C for 3 min; 8 cycles of 95 °C for 30 s, 55 °C for 30 s, and 72 °C for 30 s; and a final extension at 72 °C for 5 min. Indexed products were quantified with the Quant-iT dsDNA HS assay on a Qubit fluorometer, purified with MagSi-NGSPREP Plus magnetic beads (Steinbrenner Laborsysteme, Wiesenbach, Germany), and normalized on a Janus Automated Workstation (PerkinElmer, Waltham, MA, USA). Sequencing was performed on an Illumina MiSeq with dual indexing and the MiSeq Reagent Kit v3 (600 cycles). Mock (negative-control) samples were included for quality control.

### Sequence processing and taxonomic classification

After demultiplexing and primer trimming with Cutadapt v3.7 (Martin, 2011) reads (FASTQ) were processed in QIIME2 v2023.5 (Bolyen et al., 2019). Paired-end reads were denoised with DADA2 via q2-dada2 (Callahan et al., 2016). Reads with more than two expected errors in either direction were discarded, and chimeras were removed. Unique sequences were clustered with VSEARCH (Rognes et al., 2016) via q2-vsearch (Rideout et al., 2014) at a 99% identity cut-off against allele databases. 16S rRNA reads were classified with a V1–V3 16S feature-classifier (via q2-feature-classifier) against SILVA 138 99% OTUs (Pruesse et al., 2007). The staphylococcal amplicon scheme used a database of all *tuf2* alleles from staphylococcal genomes available in GenBank (as of January 2025). Prior to final analysis, non-bacterial reads were removed, and relative abundances were then renormalized. The filtered count data for all samples are provided in Supplementary Data, including genus-level profiles from the 16S rRNA V1–V3 analysis and *Staphylococcus* species-level profiles from the *tuf2* amplicon sequencing. After quality filtering, chimera removal, and exclusion of nonbacterial reads, the median sequencing depth of the 23 analyzed samples was 8,008 reads per sample for V1–V3 profiling (interquartile range, 6,058-9,079; range, 4,045-14,828) and 16,907 reads per sample for *tuf2* profiling (interquartile range, 11,568-24,046; range, 565-49,500). One *tuf2* library (Patient 5, Day 1) had clearly lower read depth than the remaining samples (565 reads). The sample was retained because its taxonomic profile was interpretable and consistent with the corresponding culture-based findings.

### Spatial analysis of culturable bacteria using the Bactogram method

At the time points indicated in Figure 1, the spatial distribution of culturable bacteria across the wound and surrounding skin was assessed using the Bactogram method, as previously described (Wallblom et al., 2024a). In brief, moist sterile filter paper was centered over the wound to collect cultivable bacteria with their spatial distribution retained, and the same filter paper was then used to make sequential imprints on three agar media: non-selective Todd-Hewitt agar showing total aerobic growth; the chromogenic CHROMagar *S. aureus* (CHROMagar Microbiology, Paris, France), on which presumptive *S. aureus* appears as pink/mauve colonies and other bacteria as white or blue (Carricajo et al., 2001); and the chromogenic UriSelect 4 agar (Bio-Rad, Marnes-la-Coquette, France), commonly used for differentiation and presumptive identification of urinary tract pathogens (Perry et al., 2003). The Bactogram was collected before cleaning and swabbing to avoid disrupting the surface bacteria. Plates were incubated aerobically at 37 °C in 5% CO₂ overnight (16 hours).

After incubation, plates were photographed for documentation and visual comparison. Todd-Hewitt plates were imaged on a ChemiDoc MP Imaging System with Image Lab software (Bio-Rad, Hercules, CA, USA) using the Stain Free Blot application, yielding grayscale images. CHROMagar *S. aureus* and UriSelect 4 plates, on which colony color is informative, were imaged in color with a DSLR camera (EOS Rebel T7i, Canon, Tokyo, Japan) fitted with a 60 mm f/2.8 macro lens (EF-S 60 mm, Canon). To standardize photography, the camera was mounted on a copy stand (RS 2 XA, Kaiser Fototechnik, Buchen, Germany), and the plates were photographed inside a custom-built black-plastic dark box illuminated by two LED panels (LEDP260C, Godox, Shenzhen, China). Images were assessed qualitatively.

### Statistical analysis

Given the small number of patients (n = 5), analyses were primarily descriptive without formal hypothesis testing. Bacterial loads (CFU per swab and CFU/cm² of dressing) are presented per patient and per study day on logarithmic scales, and the *S. aureus* fraction of the culturable load is reported as a proportion of total colonies as measured on CHROMagar *S. aureus*. Culture-independent data are summarized as genus-level (16S rRNA) and species-level (*tuf2*). Relative abundances, alpha-metrics, and culture-based identities (MALDI-TOF MS) were analyzed qualitatively only. Concordance between modalities was assessed qualitatively. The relationship between open wound area and bacterial load (total and *S. aureus* per swab) was evaluated visually and using Spearman’s rank correlation, p-values are reported two-sided and are presented descriptively, without adjustment for multiple comparisons or a formal significance threshold. Data handling, statistical analysis, and visualization were performed in R (R Foundation for Statistical Computing, Vienna, Austria) and GraphPad Prism (GraphPad Software, Boston, MA, USA). Expected samples that were unavailable for analysis were excluded from the corresponding analysis without imputation and are indicated in the figures. The underlying data used in the figures and analyses in this manuscript are provided in the Supplementary Data.

## Results

### Patient and wound characteristics

All five enrolled participants carried genetically confirmed pathogenic *COL7A1* variants and were classified as having severe RDEB based on genotype and clinical phenotype. All participants were male, with a median age of 18 years (range, 15–21 years), and had been diagnosed with RDEB in early childhood. All were receiving concomitant medications for RDEB, although no topical concomitant medication was applied directly to the study wounds. Routine antiseptic bathing was permitted when prescribed by the treating physician, provided that its frequency remained unchanged during the study. Patient 1 reported regular chlorine baths and local application of potassium permanganate to non-study wounds, whereas Patient 4 reported regular potassium permanganate baths. The patient demographics and concomitant medications have previously been reported in full in Wallblom et al. (2025).

The standard-of-care control wounds were located on the left upper leg in Patient 1 (approximate age, 5 weeks), right upper leg in Patient 2 (18 weeks), left lower leg in Patient 3 (5 weeks), right hip in Patient 4 (4 weeks), and right upper leg in Patient 5 (8 weeks). Wound ages were self-reported and should therefore be considered approximate.

### Wounds showed heterogeneous healing trajectories and variable bacterial autofluorescence

Three complementary imaging modalities were used to assess wound appearance, open wound area, and bacterial autofluorescence over time: clinical photography, laser-assisted digital planimetry, and handheld autofluorescence imaging (Figure 2A). The wounds exhibited heterogeneous wound-healing courses during the 4-week study (Figure 2B). At baseline, the median open area was 2.4 cm² (range 2.3–8.0 cm²). Three of the five wounds enlarged by Day 8, and two of these remained above their baseline area through Day 22. By Day 29, one wound (Patient 1) had closed completely, and three had contracted by ≥50% from baseline, whereas the wound of Patient 5 had more than doubled in area (8.0 to 19.0 cm²). Standardized photography reflected the same divergence by Day 29 (Figure 2C): Patient 1 had completely re-epithelialized, with only residual erythema, and Patient 3 showed near-complete healing with some residual crusting. Patient 2 had a newly formed blister within the original wound area. Patient 4 retained a small residual wound adjacent to larger wound areas, and Patient 5 showed an enlarging, fibrin-covered wound with a new satellite wound.

**Figure 2.**
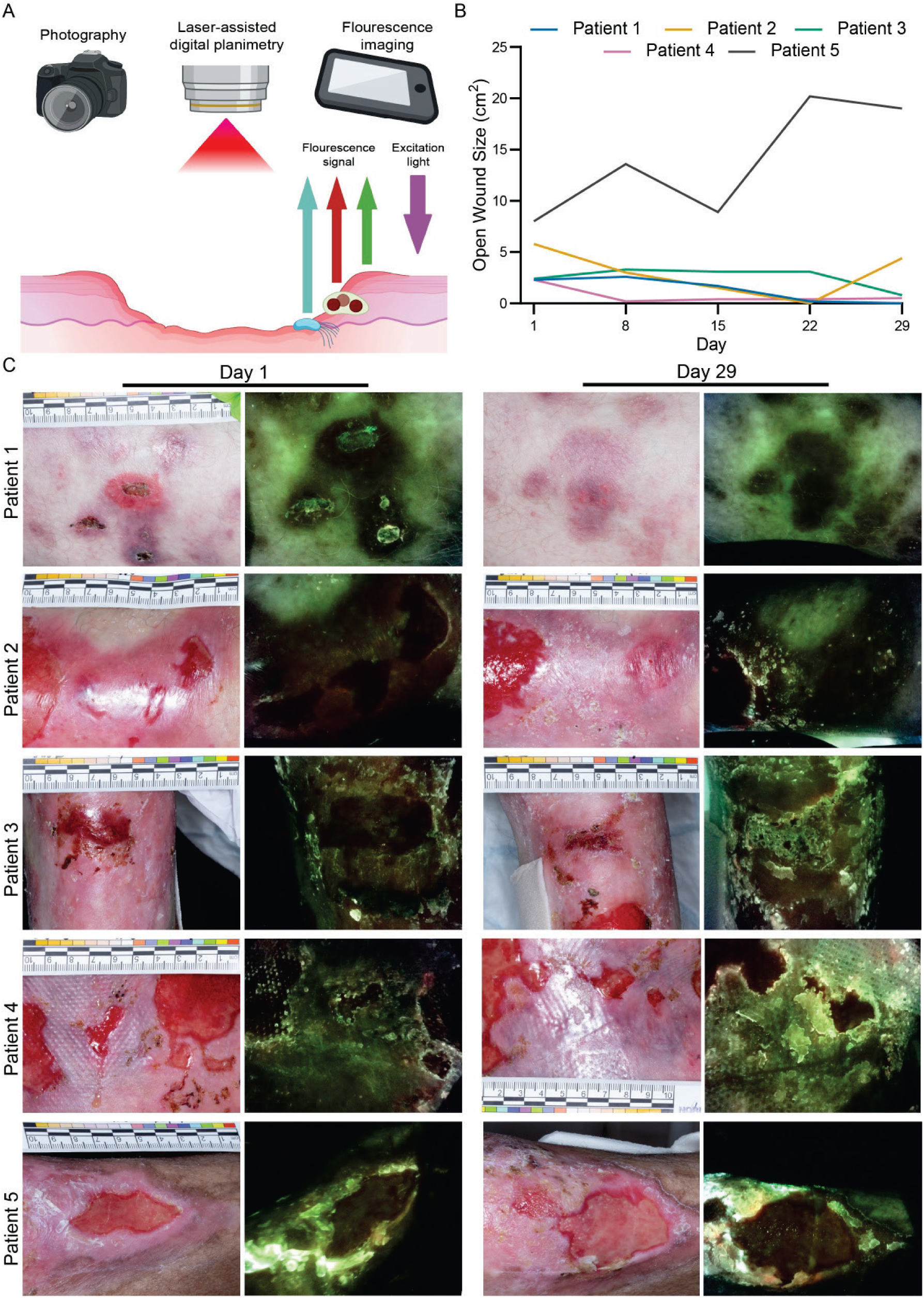
Noninvasive multimodal imaging shows heterogeneous wound-healing trajectories and variable bacterial autofluorescence in five patients with recessive dystrophic epidermolysis bullosa. (A) Three noninvasive image modalities were used: digital photography, laser-assisted digital planimetry (SilhouetteStar), and handheld autofluorescence imaging (MolecuLight i:X). The schematic shows the fluorescence principle for the MolecuLight i:X device used, with violet excitation light at 405 nm eliciting: green, tissue collagen autofluorescence; red, porphyrin- producing bacteria; cyan/white, indicate *Pseudomonas aeruginosa*. Clearly visible red or cyan bacterial fluorescence is generally associated with an elevated bacterial burden (>10^4^ CFU/g). (B) Open wound area (cm²) of the index wound as measured by laser-assisted digital planimetry at Days 1, 8, 15, 22, and 29 (one measurement per visit; Patients 1–5 by color). The measurement for Patient 2 on Day 15 was not available. (**C**) Paired standard photographs (left) and MolecuLight autofluorescence images (right) of each patient’s wound on Days 1 and 29. Patients have given written consent regarding the publication of de-identified images.

Handheld autofluorescence imaging (MolecuLight i:X) produced qualitatively interpretable bacterial signals in most patients (Figure 2C). In Patient 1, no clear bacterial fluorescence was detected at either time point. In Patient 2, red fluorescence was visible at the wound margins on Day 1 but was absent on Day 29 at the site of the newly formed blister within the original wound area. However, red fluorescence was present in the adjacent wound visible on Day 29. In Patient 3, no bacterial fluorescence was observed on Day 1, whereas a weak red signal was seen at the edge of the residual crust on Day 29. In Patient 4, red fluorescence was evident in the wound to the right of the index wound on Day 1 and at the edge of both the index and adjacent wounds on Day 29. In Patient 5, red fluorescence was not clearly visible on Day 1, but distinct white/cyan fluorescence, indicative of *P. aeruginosa*, was observed in a scaling area. By Day 29, red fluorescence appeared as distinct islands at the edge of the index wound, together with white/cyan fluorescence along several parts of the wound border.

Thus, both clinical wound status and autofluorescence patterns varied considerably among patients and over time, reflecting the dynamic nature of EB wounds. We next examined whether molecular and culture-based analyses identified a common microbiological pattern underlying these heterogeneous clinical appearances.

### Molecular and culture-based profiling identified *Staphylococcus aureus* as the predominant organism

Bacterial composition was assessed using complementary molecular and culture-based workflows, comprising 16S rRNA and staphylococcal *tuf2* amplicon sequencing of wound swabs and MALDI-TOF MS identification of colonies cultured from wound swabs and dressing fluid (Figure 3A). Genus-level alpha diversity was low, with a median observed richness of 3 genera (range, 2-5) and a Shannon index of 0.474 (range, 0.105-1.136). *Staphylococcus* was the most abundant genus in 22 of 23 samples, with a median relative abundance of 85.6% (range, 41.0-98.0%) (Figure 3B). The next most frequently detected genera were *Corynebacterium* (20 of 23 samples) and *Streptococcus* (15 of 23), followed by *Cutibacterium* (12 of 23) and *Pseudomonas* (5 of 23). These genera were usually present as minor components but occasionally reached substantial proportions, most notably *Streptococcus* in Patient 5 (36.2% and 37.9% at Days 15 and 29) and Patient 3 (22.5% at Day 22), and *Corynebacterium* in Patient 4 (32.0% at Day 15). Only modest longitudinal variations were seen, with median richness being 3 at both Days 1 and 29, whereas median Shannon diversity was 0.366 (range, 0.105-0.886) at Day 1 and 0.784 (range, 0.179-1.136) at Day 29. The main compositional exception at the genus level occurred in Patient 1 at Day 29, when the measured wound had closed, and *Cutibacterium* exceeded *Staphylococcus*.

**Figure 3.**
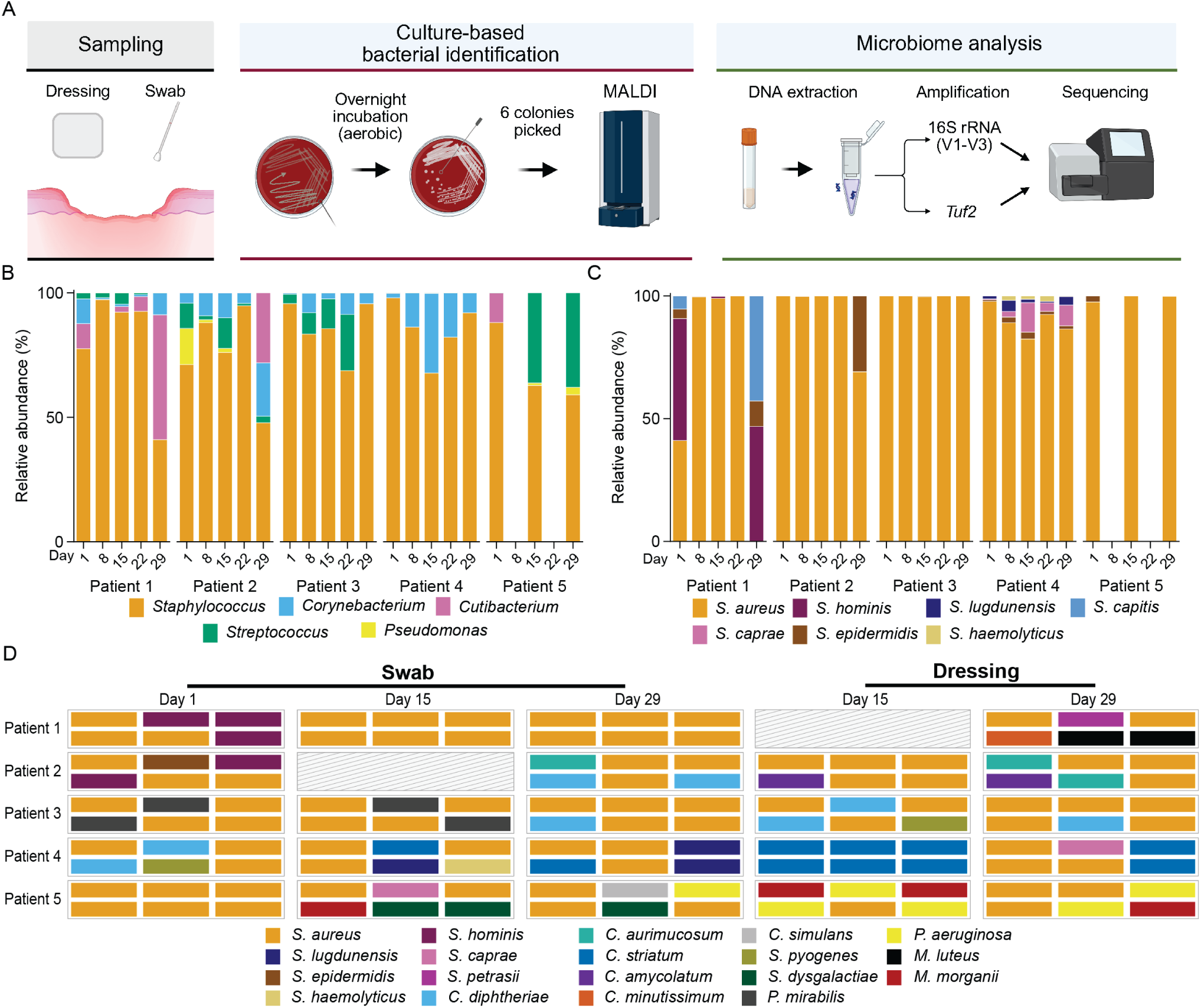
Longitudinal culture-independent and culture-based profiling shows that staphylococci, predominantly *Staphylococcus aureus*, dominate the wound bacterial microbiota in recessive dystrophic epidermolysis bullosa. (**A**) Sampling and analysis workflow. A cotton swab was used for culturing and a FLOQswab for microbiome sequencing. (**B**) Longitudinal stacked bar graph of genus-level relative abundance (%) by 16S rRNA (V1–V3) sequencing. Colors denote genera observed (key). (**C**) Longitudinal stacked bar graph of species-level relative abundance (%) of staphylococci by *tuf2* sequencing. Colors denote species observed (key). (**D**) Species-level identification of aerobically cultured bacteria using matrix-assisted laser desorption/ionization time-of-flight mass spectrometry, displayed as a categorical tile plot. Six colonies were picked per sample, from both the swab and the dressing fluid after overnight culture on blood agar. Each tile represents one colony, and colors denote the identified species according to the key. Each group of six tiles for a given patient, sampling method, and day represents the six colonies picked from that sample. Absent bars (**B, C**) and hatched cells (**D**) indicate samples not available for analysis at that visit.

Species-level staphylococcal *tuf2* profiling showed that *S. aureus* was the most abundant staphylococcal species in 21 of 23 available profiles (median relative abundance, 99.7%; range, 0-100%), indicating that the genus-level *Staphylococcus* signal detected by 16S rRNA sequencing predominantly represented *S. aureus* (Figure 3C). The most notable exception was seen in Patient 1, where *S. aureus* was co-dominant with *S. hominis* at Day 1, shifting to *S. aureus* dominance at Days 8–22 and then to dominance by coagulase-negative staphylococci (*S. capitis, S. hominis*, and *S. epidermidis*) at the Day 29 time point, when the wound had completely healed. Among the remaining staphylococcal species, *S. epidermidis* was the most common coagulase-negative species overall (present in 14 of 23 profiles; up to 30.9% in Patient 2 at Day 29), whereas *S. caprae*, *S. lugdunensis*, and *S. haemolyticus* occurred almost exclusively in Patient 4 (*S. caprae* reaching 12.0% at Day 15) and *S. hominis* and *S. capitis* only in Patient 1. Across all samples, the staphylococcal diversity was generally low, with a median observed species richness of 2 (range, 1-5) and a median Shannon index of 0.020 (range, 0-0.995). Only modest longitudinal variations were observed, with median richness at 2 on both Days 1 and 29, and median Shannon diversity at 0.116 (range, 0-0.995) and 0.525 (range, 0-0.954), respectively.

MALDI-TOF MS confirmed that viable *S. aureus* was frequently recovered from both swab and dressing-fluid cultures (Figure 3D). Among the six selected colonies per sample, *S. aureus* was the most frequent or co-most frequent species in all 14 available swab cultures and in seven of nine available dressing-fluid cultures. The two main discordances between the swab and the paired dressing fluid occurred on Day 15: in Patient 4, the swab yielded predominantly *S. aureus*, whereas all six dressing-fluid colonies were *Corynebacterium striatum*; and in Patient 5, the swab yielded *S. aureus* co-dominant with *Streptococcus dysgalactiae*, whereas the dressing fluid was dominated by *P. aeruginosa* and *Morganella morganii*. Other intermittently recovered organisms included coagulase-negative staphylococci, corynebacteria, streptococci, *Proteus mirabilis*, and *P. aeruginosa*.

Having established that *S. aureus* dominated both the molecular profiles and the viable cultured population, we next determined whether this predominance also represented the major quantitative bacterial burden.

### Viable aerobic bacterial loads varied substantially without apparent association with open wound area

To quantify viable aerobic bacterial burden and the proportion attributable to presumptive *S. aureus*, wound-swab and dressing samples were serially diluted and cultured in parallel on Todd-Hewitt agar and CHROMagar *S. aureus* (Figure 4A). On non-selective Todd-Hewitt agar, swab-derived loads had a median of 2.5×10⁴ CFU/swab and spanned more than four orders of magnitude across samples (1×10² to 3.2×10⁶ CFU/swab) (Figure 4B). Dressing-derived loads, normalized to sampled area, were generally higher (median 7.7×10⁵ CFU/cm²; range 8.3 × 10¹ – 1.88 × 10^8^ CFU/cm²) (Figure 4C). Patient 5 carried the highest dressing burdens (∼10⁷–10⁸ CFU/cm²), whereas Patient 1 had the lowest, falling below 10² CFU/cm² by Day 29 as the wound closed.

**Figure 4.**
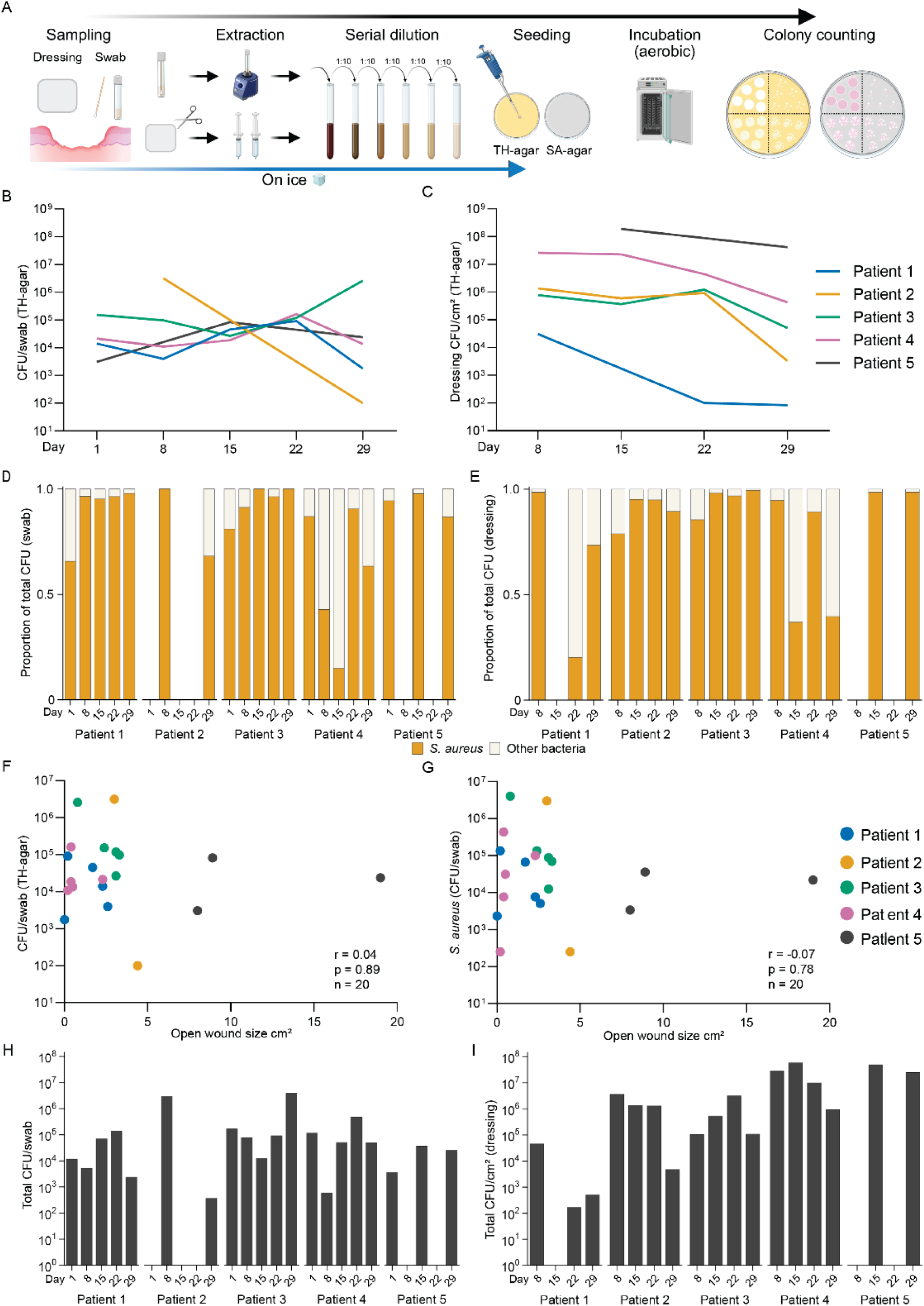
Aerobic quantitative culture shows a generally high but heterogeneous, *Staphylococcus aureus-*dominated viable bacterial load in recessive dystrophic epidermolysis bullosa wounds in recessive dystrophic epidermolysis bullosa wounds, without apparent association with open wound area. (A) Workflow for quantitative aerobic culture of wound swabs and wound dressings. (**B, C**) Longitudinal total culturable aerobic bacterial load on Todd-Hewitt agar from wound swabs (B) and wound dressings (**C**). Swab-associated load is expressed as CFU/swab, whereas dressing-associated load is normalized to sampled dressing area and expressed as CFU/cm² dressing. (**D, E**) Proportion of colony counts on CHROMagar *S. aureus* agar classified by colony color in swab (**D**) and dressing (**E**) samples over time. For each sample, pink colonies were counted as presumptive *S. aureus*, whereas non-pink colonies were classified as other bacteria. Stacked bars show these categories as fractions of the total CFU recovered on CHROMagar *S. aureus* agar from that sample. (**F, G**) Relationship between open wound area and aerobic bacterial burden. Open wound area was compared with total CFU/swab on Todd- Hewitt agar (**F**) and presumptive *S. aureus* CFU/swab on CHROMagar *S. aureus* agar (**G**). Each point represents one sample and is colored by patient. Spearman correlation coefficients, p-values, and sample sizes are shown in each panel. (**H, I**) Longitudinal total bacterial load on CHROMagar *S. aureus* agar from swabs (**H**) and dressings (**I**). Counts include all colonies growing on CHROMagar *S. aureus*. All bacterial loads are shown on logarithmic scales. Missing bars or interrupted line segments indicate samples that were not available for analysis at the corresponding visit.

Quantitative aerobic culture on chromogenic CHROMagar *S. aureus* indicated that this load was predominantly *S. aureus*. Presumptive *S. aureus* (pink colonies) constituted a median of 93% of the recoverable swab load (range 15–100%) and 95% of the dressing load (range 20– 99%) (Figure 4D and 4E), and the corresponding total chromogenic counts largely tracked the Todd-Hewitt data (Figure 4H and 4I).

Thus, the strong predominance *of S. aureus* identified by *tuf2* sequencing and MALDI-TOF MS was also reflected quantitatively: colonies with a phenotype compatible with *S. aureus* accounted for most of the viable aerobic burden in both swab and dressing samples.

Bacterial burden was not clearly associated with clinical wound size. Open wound area showed no correlation with total swab bacterial load (Spearman r = 0.04, p = 0.89; Figure 4F) or with presumptive *S. aureus* swab bacterial load (r = −0.07, p = 0.78; Figure 4G). These exploratory pooled correlations should be interpreted cautiously because repeated observations were obtained from only five patients. Nevertheless, the longitudinal patterns similarly showed that wound size reduction was not consistently accompanied by a reduction in viable bacterial burden. High *S. aureus* burdens were recovered from small, nearly re-epithelialized wounds, for example, ∼3×10⁶ CFU/swab from Patient 3 at Day 29 (open area 0.8 cm²) and 10⁵ CFU/swab from Patient 1 at Day 22 (0.2 cm²). However, for the only completely re-epithelialized wound (Patient 1, Day 29), the culturable load was lower (∼2×10³ CFU/swab).

Collectively, these observations indicate that marked wound reduction and near-complete re-epithelialization did not necessarily correspond to microbiological clearance. We therefore used spatial imprint culturing to determine how the persistent viable, *S. aureus-*dominated population was distributed across the wound and surrounding skin.

### Bactogram spatial imprinting mapped viable *Staphylococcus aureus*-dominated growth across the wound and surrounding skin

Given the consistent *S. aureus* predominance demonstrated by sequencing, MALDI-TOF MS, and quantitative chromogenic culture, we investigated whether Bactogram imprinting could provide a spatially resolved representation of this viable bacterial population. A single imprint was sequentially transferred onto Todd-Hewitt, CHROMagar *S. aureus*, and UriSelect 4 agar (Figure 5A). Bactograms demonstrated growth across both wound and peri-wound regions in all five patients at Days 1 and 29 (Figure 5B).

**Figure 5.**
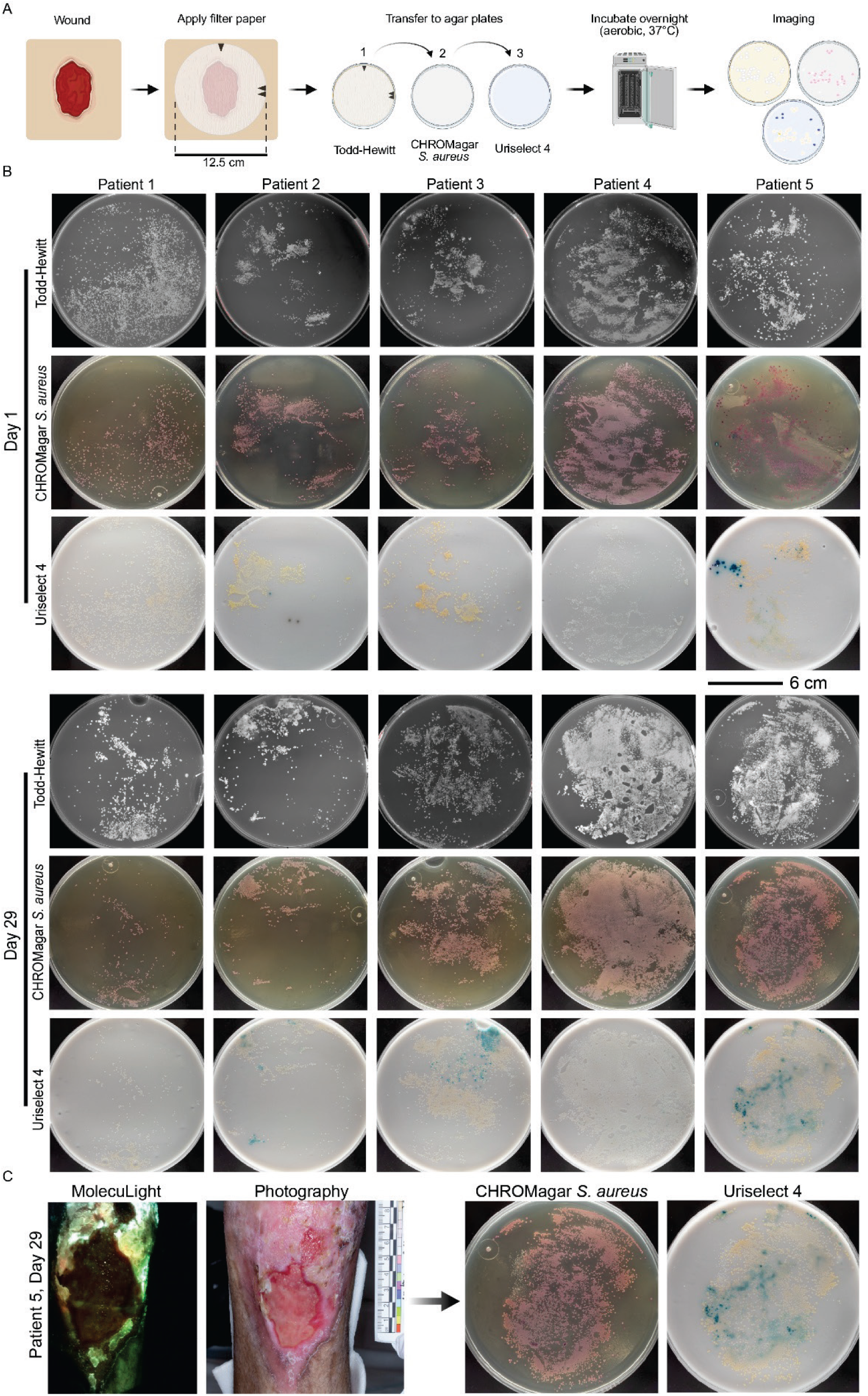
Spatial analysis of aerobic culturable bacteria using the Bactogram method suggests a spatially and longitudinally *Staphylococcus aureus*-dominant microbiota in wounds and surrounding skin of recessive dystrophic epidermolysis bullosa. (**A**) Bactogram workflow. A sterile filter paper was centered on the wound and surrounding skin and sequentially transferred to Todd-Hewitt agar, CHROMagar *S. aureus*, and UriSelect 4 before overnight aerobic incubation. Todd-Hewitt agar shows total aerobic growth, whereas pink/mauve colonies on CHROMagar *S. aureus* are compatible with presumptive *S. aureus.* UriSelect 4, used as a non-selective chromogenic medium for visualization of common urinary tract pathogens, was applied here for exploratory visualization of selected wound bacteria. Expected color reactions include pink colonies compatible with *E. coli*, turquoise-blue colonies compatible with enterococci, blue-purple colonies compatible with the *Klebsiella– Enterobacter–Serratia–Citrobacter* group, and orange-brown colonies with a brownish halo compatible with the *Proteus–Providencia–Morganella* group. (**B**) Bactograms from all five participants on Days 1 and 29. Corresponding plates are shown for each participant and visit, with growth assessed qualitatively. (**C**) Illustrative case from Patient 5 at Day 29 integrating fluorescence imaging and clinical photography. The clinical image was visually aligned and size-matched to the Bactogram imprints to illustrate the approximate spatial correspondence.

Growth on CHROMagar *S. aureus* broadly overlapped the regions of aerobic growth on Todd-Hewitt agar, and pink/mauve colonies indicative of *S. aureus* dominated most imprints. This was consistent with the *S. aureus*-dominant bacterial microbiota identified through sequencing and MALDI-TOF MS analyses of swab and dressing samples. The spatial extent and density of visible growth varied among participants and over time: growth was qualitatively reduced in Patient 1 at Day 29, whereas extensive growth persisted in Patients 3–5 despite reductions in measured wound area in Patients 3 and 4, mirroring the apparent lack of association between bacterial burden and open wound area. UriSelect 4 imprints showed localized chromogenic variations within otherwise predominantly pale (white to white-yellow) colonies, consistent with a mainly staphylococcal bacterial microbiota. Blue- or turquoise-toned colonies were visible in Patients 1 and 5 at Day 1 and in Patients 2, 3, and 5 at Day 29, indicating spatially restricted, non-*S. aureus* colony phenotypes.

Patient 5 at Day 29 illustrates the complementary information provided by the different modalities (Figure 5C). The clinical image was visually aligned and size-matched to the corresponding culture imprints. Pink/mauve colonies occupied much of the CHROMagar *S. aureus* imprint, whereas blue/turquoise phenotypes on UriSelect 4 were more localized. At the same visit, aerobic loads were 2.38×10⁴ CFU/swab and 4.15×10⁷ CFU/cm² dressing, of which 86.8% and 98.6%, respectively, were presumptive *S. aureus* on CHROMagar. Genus-level sequencing identified *Staphylococcus* (59.1%), *Streptococcus* (37.9%), and *Pseudomonas* (3.0%), whereas 99.9% of *tuf2* reads were assigned to *S. aureus*. MALDI-TOF MS recovered *S. aureus* and *P. aeruginosa* from both swab and dressing-fluid cultures; *S. dysgalactiae* and *C. simulans* were recovered only from the swab, and *M. morganii* only from dressing fluid.

Together, these readouts characterize an enlarging wound with a high burden of aerobic, viable bacteria and a polymicrobial, spatially heterogeneous microbiota dominated by *S. aureus.* No single modality captured all dimensions of this microbiology: sequencing defined bacterial composition, quantitative culture measured viable burden, MALDI-TOF MS identified cultivable species, autofluorescence provided an immediate but variably detectable signal, and the Bactogram retained the spatial distribution of viable bacterial growth.

## Discussion

This exploratory longitudinal study combined 16S rRNA (V1–V3) and staphylococcal *tuf2* amplicon sequencing with culture-based species identification, spatial imprint analysis, and bacterial autofluorescence imaging to characterize standard-of-care-treated RDEB wounds from five patients over four weeks. Despite markedly different healing trajectories, the wounds shared a microbiological pattern of low-diversity, *Staphylococcus*-dominated communities in which *S. aureus* accounted for most of the staphylococcal signal and most of the recoverable aerobic burden. Viable aerobic bacterial loads were generally high and varied by several orders of magnitude, and were not clearly associated with the open wound area. The modalities converged at the cohort level but provided distinct information on bacterial composition, viability, abundance, and spatial distribution in RDEB wounds.

The dominance of staphylococci, and *S. aureus* in particular, in EB wounds is well established, both in culture-based surveys (Brandling-Bennett and Morel, 2010; Fuentes et al., 2023; Levin et al., 2021; Singer et al., 2018) and in sequencing studies (Amsalu et al., 2026; Bar et al., 2021; Fuentes et al., 2018; Horev et al., 2023; Reimer-Taschenbrecker et al., 2022). The present study extends these observations by demonstrating close cohort-level correspondence between broad community profiling, staphylococcal species resolution, viable-isolate identification, quantitative culture, and spatial imprinting. A specific contribution was the use of *tuf2* amplicon sequencing to reliably resolve the staphylococcal component of the wound community in EB (Ahle et al., 2021; Feidenhansl et al., 2024). Most EB sequencing studies have used 16S rRNA profiling (Amsalu et al., 2026; Bar et al., 2021; Horev et al., 2023; Reimer-Taschenbrecker et al., 2022), which reliably identifies staphylococci only at the genus level (Ahle et al., 2021), necessitating resource-intensive shotgun metagenomics for species-level identification, as done in Fuentes et al. (2018). Using *tuf2* amplicon sequencing, we found that the staphylococcal population was almost entirely composed of *S. aureus*, concordant with the MALDI-TOF MS identification of cultured isolates from the same wounds. The main discordance occurred in the only fully re-epithelialized wound, where *tuf2* sequencing detected no *S. aureus* despite its recovery by culture. This most likely reflects the difficulty of profiling low-biomass samples, which are sensitive to minor sampling variation and in which a low-abundance organism can be missed by a relative-abundance sequencing readout, yet still be recovered by culture (Misic et al., 2014; Ren et al., 2021; Tettamanti Boshier et al., 2020). *Tuf2* sequencing provides an intermediate approach between genus-level 16S rRNA profiling and comprehensive metagenomic or whole-genome techniques, making it well-suited for species determination of the EB-associated bacterial microbiota dominated by staphylococci.

Quantitative culture added an aerobic, viable-load dimension that sequencing cannot provide. By using both non-selective and CHROMagar *S. aureus* media, we measured both the total aerobic burden and the viable fraction attributable to presumptive *S. aureus* on the same samples. This contrasts with the relative abundances obtained by amplicon sequencing, which also include nonviable organisms and reflect composition rather than absolute abundance (Misic et al., 2014; Tettamanti Boshier et al., 2020). Aerobic viable burdens were generally high and, in most wounds, persisted as wounds contracted and re-epithelialized, with small, nearly re-epithelialized wounds still harboring high *S. aureus* loads. This is consistent with recent findings in epidermal wounds of healthy volunteers, where surface loads rose to about 10⁶ CFU per swab and remained at that level during re-epithelialization (Lundgren et al., 2025). In DEB, this tendency is likely reinforced by disease-specific factors. The skin carries persistent structural and immunological barrier defects (Cianfarani et al., 2017; Nystrom et al., 2021), including extracellular matrix–dependent impairment of antibacterial immunity (Nystrom et al., 2018) and chronic immune dysregulation at the wound (Huitema et al., 2021), which plausibly contribute to persistent bacterial colonization despite wound contraction. Even so, this colonization is not necessarily benign. *S. aureus* has been associated with disease activity (Reimer-Taschenbrecker et al., 2022) and itch (Deng et al., 2023), with emerging evidence indicating that *S. aureus* may shape the local inflammatory response in RDEB in a strain-specific manner (Jamet et al., 2025). Establishing mechanisms linking colonization to local and systemic inflammation and wound healing will require studies that pair microbiological and host-response measurements with longitudinal clinical outcomes.

A single swab usually samples only one region, with each additional swab adding contact with fragile, painful skin and requiring separate processing. van der Kooi-Pol et al. showed that colonization in EB is spatially structured, resolving distinct *S. aureus* types as separate micro- colonies within single EB wounds (van der Kooi-Pol et al., 2013). The Bactogram extends this view to the whole wound and surrounding skin at the level of aerobic viable burden and presumptive species, showing a broadly *S. aureus*–dominant surface punctuated by spatially restricted, non–*S. aureus* phenotypes (Wallblom et al., 2024a). Noninvasive autofluorescence imaging provided a rapid, contact-free view, with localized fluorescence signals suggesting a high bacterial burden, with red signals indicating porphyrin-producing bacteria and cyan/white signals indicating *P. aeruginosa* (Le et al., 2021). However, fluorescence is a qualitative readout with limited sensitivity: the signal depends on bacterial fluorophore production, burden, interference by blood, and depth. Substantial culturable loads were sometimes present without a visible signal, so a negative signal does not exclude high colonization. Despite this, a growing body of evidence in chronic, non-EB wounds suggests that bacterial autofluorescence imaging can guide clinicians to the areas of highest bacterial burden, helping to target both wound sampling and interventions such as debridement (Badrie et al., 2025; Le et al., 2021).

To our knowledge, the two spatial methods used here, the Bactogram and autofluorescence imaging, were applied in RDEB for the first time. Beyond these, used dressings provided an easy-to-collect sample that yielded substantial viable bacterial populations while integrating a larger surface area and a longer exposure window than a single swab. Because dressing selection and handling are major challenges in EB care (Bruckner et al., 2020; Denyer et al., 2017), and because occlusive dressings can accumulate high bacterial loads (Hutchinson and Lawrence, 1991; Scheer et al., 2024) that also contribute to wound malodor (Akhmetova et al., 2016; Stuermer et al., 2025), dressing-derived cultures could offer an objective readout for comparing wound-care materials or wear protocols. More broadly, minimally invasive approaches such as autofluorescence imaging, the Bactogram, and dressing sampling are well-suited to fragile EB skin and could be incorporated into future studies and clinical trials as exploratory readouts for longitudinal monitoring, complementing rather than replacing conventional microbiological analysis.

This exploratory study is limited by its small, all-male cohort, which is underpowered for formal hypothesis testing, a common challenge in rare diseases such as EB (Mellerio, 2022; Prodinger et al., 2020). Only one wound closed completely, and no unwounded RDEB or healthy-skin controls were included. Some samples were unavailable at individual visits or for specific modalities, reducing the completeness of the longitudinal data. However, because every patient still contributed repeated measurements, these gaps are unlikely to have meaningfully affected our main conclusions. Several methods were qualitative, and all measurements concerned the wound surface; deeper tissue, biofilm architecture, strain identity, antimicrobial resistance, and host inflammation were considered beyond the scope of the study. The results should consequently be regarded as primarily descriptive and hypothesis-generating. The strength of the study lies instead in its longitudinal assessment of untreated control wounds under a standardized dressing and cleaning protocol, using several methods with distinct measurement principles that converged on the same main finding while each added complementary detail.

In short, the multimodal microbiological assessment converged on *S. aureus* as the dominant bacterium in RDEB wounds and adjacent skin, with high aerobic viable loads that showed no apparent association with wound size and often persisted as wounds contracted. We suggest that bacterial burden, composition, and spatial distribution should be considered as distinct, complementary dimensions when assessing wound microbiology and designing trials in EB. Larger longitudinal cohorts linked to clinical outcomes are needed to determine which methods provide the greatest clinical and translational value.

## Acknowledgments

The data presented here was supported by grants from the Swedish Research Council (2020- 02016, 2025-02401), Edvard Welanders Stiftelse and Finsenstiftelsen (Hudfonden), the Österlund Foundations, and the Swedish Government Funds for Clinical Research (ALF). Xinnate AB provided the project management resources and expertise for the regulatory development enabling the clinical parts of the Safety study that generated the control samples and data used in this work. We thank the research team at the Biomedical Center, Lund University, for their invaluable support with sample handling and microbiological assays: Francisco Cardoso, Congyu Luo, and Ann-Charlotte Strömdahl. We acknowledge the support of research nurses Ulrike Spetz-Nyström and Maria Wall, as well as other staff at Clinical Trial Consultants AB’s Oscar Unit. We are thankful for the support of Kerstin Persson and Emma Belfrage at the Department of Dermatology in Lund, as well as Maria Wass at the Department of Dermatology at Uppsala University Hospital.

## Ethics Statement

The study was conducted in accordance with the latest revision of the Declaration of Helsinki and the International Council for Harmonisation E6 Good Clinical Practice guideline, following approval by the Swedish Medical Products Agency and the Swedish Ethical Review Authority (Etikprövningsmyndigheten; application no. 2022–00527-01; amendment no. 2023–05051-02). Written informed consent was obtained from all participants or, for those under 18 years of age, from their parents or legal guardians before study participation. Patients have given written consent regarding the publication of de-identified images.

## Data Availability Statement

Additional data supporting the findings of this study are available from the corresponding author upon reasonable request, except for information that cannot be shared publicly for legal reasons, as it could compromise the privacy of research participants. The underlying data used in the figures and analyses in this manuscript are provided in the Supplementary Data.

## Author Contributions

The following authors contributed to each of the following roles (defined according to the CRediT taxonomy): Conceptualization and design of methodology, KH, ES, HB, AS, KW; formal analysis (statistics), KW; investigation (performing experiments and data collection) and validation, KH, BN, EH, HB, KW; resources (provision of patients and samples), KH, ES, SL, TH, KW; data curation, BN, EH, HB, AS, KW; writing (original draft preparation), AS, KW; writing (review and editing), KH, ES, SL, TH, BN, EH, HB, AS, KW; visualization, KW; supervision, AS, KW; project administration, AS, KW; funding acquisition, AS. All authors have read and agreed to the published version of the manuscript.

